# Late-onset unexplained epilepsy as a risk factor for cognitive impairment and dementia: Protocol for a multi-center prospective longitudinal observational study (ELUCID)

**DOI:** 10.1101/2025.06.16.25329698

**Authors:** Alice D. Lam, Emily L. Johnson, Rani A. Sarkis, Leah J. Blank, Tyler E. Gaston, Mouhsin M. Shafi, Rodrigo Zepeda, Kyle R. Pellerin, Nathalie Jette, Douglas N. Greve, Lori B. Chibnik, Rebecca E. Amariglio, Gad A. Marshall, M. Brandon Westover

## Abstract

**Background:** Late-onset unexplained epilepsy (LoUE), defined as epilepsy onset after age 55 without an obvious cause, is an important risk factor for dementia. Studies have shown that 10-25% of individuals with LoUE develop dementia within three to four years following their first seizure. However, the mechanisms underlying progression from LoUE to dementia remain poorly understood. The goals of the ELUCID study are to identify risk factors associated with development of cognitive decline and dementia in LoUE, and to develop tools to identify patients at high risk for these outcomes and thereby establish a foundation for dementia prevention strategies in this population.

**Methods and analysis:** ELUCID is a multi-center prospective longitudinal observational study that will enroll 600 participants aged 55 or older with LoUE across seven U.S. medical centers. Participants undergo a baseline evaluation that includes a detailed clinical history, cognitive testing, brain MRI, overnight scalp EEG, and blood biomarkers. Participants will be followed at six-month intervals to record cognitive and neurological changes, with the primary outcomes of interest being development of mild cognitive impairment and/or dementia. This study aims to establish LoUE disease subtypes based on biomarkers, cognitive trajectories, and imaging features, and to develop a risk stratification tool for predicting risk for cognitive decline and dementia in patients presenting with LoUE.

**Ethics and Dissemination:** ELUCID has obtained IRB approval (# 2023P001566, August 2023), with the MassGeneral Brigham IRB serving as the single IRB of record. All de-identified study data will be made publicly available on completion of the study.

## INTRODUCTION

### Background and rationale

#### Late-onset unexplained epilepsy (LoUE) is a growing public health priority

LoUE, defined here as epilepsy onset after age 55 in which no clear etiology can be identified, accounts for up to 25% of all new cases of epilepsy and afflicts 50,000 new patients in the U.S. each year^1–10^. The impact of LoUE will further increase in upcoming decades, as: 1) Older adults are the fastest growing demographic in the U.S, and are expected to double in size over the next 30-40 years^11^; and 2) The incidence rate of epilepsy in older adults is rising^1,9,12^. The incidence rate of epilepsy in adults aged 65 and older doubled in Rochester, MN between 1935-1984^1^; and quadrupled in Finland between 1973-2013^9,12^. This increase is not attributable to better ascertainment of epilepsy cases, as studies reporting these increases reported a decrease or no change in epilepsy incidence in children and younger adults in the same period^1,9,12^. Moreover, the incidence rates of the 3 most common causes of epilepsy in older adults (stroke, dementia, traumatic brain injury) have all decreased in recent decades^1,13–22^. Much of the growing incidence rate of epilepsy in older adults thus remains unexplained and falls within the category of LoUE.

#### Increased risk of dementia, stroke, and mortality in LoUE

While current clinical management of LoUE focuses primarily on preventing seizures, excess morbidity and mortality from LoUE are more closely related to development of dementia and stroke than to seizures^23^. LoUE is considered to be highly pharmacosensitive. Over 80% of individuals with LoUE achieve seizure-freedom with medications alone, and seizures account for < 1% of all deaths in LoUE^23–29^. Yet, in epidemiologic studies, individuals with LoUE have a 2-3-fold increased risk for developing dementia^30–34^ and a 2-3-fold increased risk for developing stroke^35^, compared to similar people without epilepsy. LoUE is also associated with a 3-fold increased risk of mortality, with excess risk of dementia and stroke being the major contributors^23,36^. A reassessment of our clinical approach to LoUE is needed. Development of LoUE may be a critical point for early intervention to prevent subsequent dementia or stroke. Understanding the mechanisms that lead to dementia and stroke in LoUE will be critical for developing therapies to forestall and prevent these outcomes. These mechanisms may also inform novel and shared pathways for treating other types of dementia.

#### Need for a precision medicine approach to preventing dementia and stroke in LoUE

We currently have no clear understanding of the mechanisms underlying dementia or stroke in LoUE. Moreover, we have no tools to identify which individuals with LoUE are at greatest risk for developing these outcomes, and no treatments to prevent these outcomes in individuals with LoUE. A major shortcoming of our current clinical approach to LoUE is that LoUE is treated as a homogenous condition. Yet, substantial clinical heterogeneity exists within LoUE, and multiple pathologies are likely to underlie LoUE and its related risks of dementia and stroke. There is an urgent need to develop a precision medicine approach to preventing these outcomes in LoUE, where disease subtyping specifically informs a patient’s risk for dementia (or stroke) and guides targeted therapy to reduce this risk.

The ELUCID study will focus on primary outcomes of mild cognitive impairment and dementia, and will also track secondary outcomes including stroke, cardiovascular disease, and mortality.

### Aims of the Study

1. Establish an unbiased framework for identifying mechanisms of dementia in LoUE. We will use unsupervised machine learning approaches to organize LoUE into disease subtypes based on pathology, clinical phenotype, and cognitive trajectory.
2. Identify clinical features and biomarkers associated with development of cognitive decline and dementia in LoUE.
3. Develop practical clinical tools to forecast an individual’s risk of developing mild cognitive impairment and/or dementia after presentation with LoUE.

### Study Hypotheses

Our central hypothesis is that rigorous phenotyping of LoUE based on clinical features, comorbidities, biomarkers, and cognitive outcomes will improve mechanistic understanding of dementia (and stroke) in LoUE and accelerate development of targeted therapies to reduce risk of these outcomes in LoUE. As most individuals with LoUE do not develop these outcomes, we further hypothesize that mechanisms underlying cognitive decline or stroke in LoUE will be distinct from those that cause seizures alone (without cognitive decline or stroke). Defining these mechanisms will inform therapeutic approaches to prevent dementia and stroke in LoUE and may also inform approaches for primary prevention of LoUE.

## METHODS

### Study Design

ELUCID (Epilepsy of Late-onset Unknown etiology as a risk factor for Cognitive Impairment and Dementia) is a 7-center, prospective, longitudinal observational study focused on understanding mechanisms and predicting primary outcomes of cognitive decline and dementia in LoUE. The study will enroll 600 participants with LoUE across 7 study sites over the first 3 years of the study. ELUCID study sites are:

- Massachusetts General Hospital, Boston, MA (coordinating site)
- Beth Israel Deaconess Medical Center, Boston, MA
- Brigham and Women’s Hospital, Boston, MA
- Icahn School of Medicine at Mount Sinai, New York, NY
- Johns Hopkins Medicine, Baltimore, MD
- University of Alabama at Birmingham, Birmingham, AL
- University of Texas Southwestern, Dallas, TX

We will acquire a baseline dataset that includes detailed clinical history, brain MRI, overnight scalp EEG, blood biomarkers of neuropathology, vascular health, and inflammation, and banked plasma / DNA samples.

Longitudinally, we will track cognitive changes with annual neuropsychological testing and assess interval changes in seizure and medical history and development of primary outcomes (mild cognitive impairment and dementia) and secondary outcomes (stroke, transient ischemic attack, myocardial infarction, serious cardiac arrhythmia, and death) of interest.

### Study Population and Eligibility Criteria

The inclusion and exclusion criteria for this study are provided in **Table 1**. Participants will have experienced their first seizure after age 55 and within 3 years of enrollment. Our rationale for including individuals with first seizure is that the probability of recurrence after a first seizure in older adults is sufficiently high to meet ILAE criteria for epilepsy^37,38^. As seizures in older adults often occur with subtle symptoms^16,39,40^, we have defined formal criteria for probable and definite seizure or epilepsy for use in this study, as shown in **Table 2**. Study participants are strongly encouraged, but not required, to have a study partner who knows them well and who can accompany them to study visits to provide collateral information on seizure frequency and daily functioning.

**Table 1.** ELUCID Inclusion and Exclusion Criteria.

| Inclusion Criteria | Exclusion Criteria |
| --- | --- |
| • Age $\geq 55$ at time of first seizure<br>• First seizure occurred within 3 years of enrollment | • History of dementia or intellectual disability<br>• Identifiable etiology for seizure (e.g., cortical stroke, hemorrhage, severe traumatic brain injury, tumor)<br>• Provoked seizures (e.g., metabolic, substance induced, alcohol withdrawal, acute brain injury)<br>• Prior brain surgery (including neuromodulatory devices)<br>• History of limbic encephalitis and/or current treatment with immunomodulatory maintenance therapy for presumed auto-immune epilepsy, regardless of autoantibody status<br>• Structural neocortical lesion on MRI that is clinically deemed to be an epileptogenic focus. *The following will NOT be considered exclusionary:* small vessel ischemic disease (any severity), including white matter hyperintensities and subclinical lacunar infarcts; hippocampal sclerosis; diffuse/focal cortical atrophy of unknown etiology (i.e., not related to prior injury). Decisions to exclude other incidental MRI findings that arise will be made by consensus of the study investigators.<br>• Language or hearing impairment that limits ability to participate in cognitive testing or study phone calls.<br>• Non-English speaking, or level of English proficiency at a level that would adversely impact neuropsychological assessment in English<br>• Active alcohol or substance use that may interfere with study participation or compromise study results<br>• Insufficient clinical data to rule out the above exclusion criteria |

**Table 2.** Diagnostic criteria for late-onset unexplained seizure and epilepsy.

|  | **Probable** | **Definite** |
| --- | --- | --- |
| **Seizure \* #** | Unwitnessed episode involving loss of consciousness, tongue bite, incontinence, and/or other significant injury, suggestive of generalized convulsion<br>Single transient neurologic event suggestive of focal seizure (e.g., unresponsive with automatisms; post-ictal confusion) | Witnessed generalized convulsion |
| **Epilepsy \*** | Either of the above criteria for “Probable Seizure”, in addition to EEG with epileptiform abnormality<br>Recurrent, stereotyped neurologic events, but EEG without epileptiform abnormality | 2 or more witnessed generalized convulsions separated by $\geq 24$ hours<br>1 witnessed generalized convulsion AND EEG with epileptiform abnormality<br>1 witnessed generalized convulsion AND recurrent, stereotyped neurologic events<br>Recurrent, stereotyped neurologic events AND EEG with epileptiform abnormality<br>Recurrent stereotyped neurologic events that resolve completely with anti-seizure medication |
| * Events should NOT be better explained by an etiology other than seizure |  |  |
| # Seizure as defined here is meant for individuals who have had a single event (and does not consider the fact that the high rate of seizure recurrence in this age group would qualify these individuals as having epilepsy, based on ILAE criteria) |  |  |

As noted in **Table 1**, individuals with a prior diagnosis of dementia will be excluded from the study. We will also formally assess for dementia as part of our phone screening procedure, as described below.

### Recruitment, Screening, and Enrollment

Potential participants will be identified by reviewing lists of patients seen in outpatient epilepsy clinics, epilepsy monitoring units, emergency rooms, and other clinics at study sites where a diagnosis code of epilepsy or seizure is used. We will also recruit potential participants referred by neurologists at local hospitals. We will review medical records to identify potential participants who initially satisfy our inclusion and exclusion criteria. Research coordinators at each site will contact potential participants by phone and administer a standardized questionnaire to formally assess inclusion/exclusion criteria.

During the phone screen, we will formally assess for dementia as follows. First, potential participants are assessed for subjective cognitive decline with the following question: “Have you experienced a change in your memory in the last 1-3 years?”. If they answer yes, they are asked two additional questions: “Has this been a consistent change over the last 6 months?” and “Are you concerned about this change?”. If they answer yes to either question, the research coordinator will carry out a Telephone Interview for Cognitive Status (TICS) questionnaire with the potential participant. If the TICS score indicates cognitive impairment (TICS < 31, or < 29 for those with less than a high school education) the research coordinator will ask the potential participant to identify an informant (a close family member or friend) who can provide details about their daily activities. The informant will then complete a Quick Dementia Rating System (QDRS) questionnaire for the potential participant. A QDRS score ≥ 6 indicates dementia and would exclude the potential participant from the study.

If the potential participant is unable to identify an informant, they would also be excluded from the study.

If the potential participant qualifies for the study and is interested in participating, written informed consent will be obtained prior to enrollment.

### Study Visits and Procedures

All ELUCID participants undergo a Baseline Assessment that includes a detailed clinical history, physical exam, neuropsychological assessment, 3T Brain MRI, overnight scalp EEG, and blood draw. After the Baseline Assessment, study visits occur every 6 months, alternating as Phone Visits and In-Person Visits, described further below. A schematic of the study protocol is shown in **Figure 1**.

**Figure 1.**
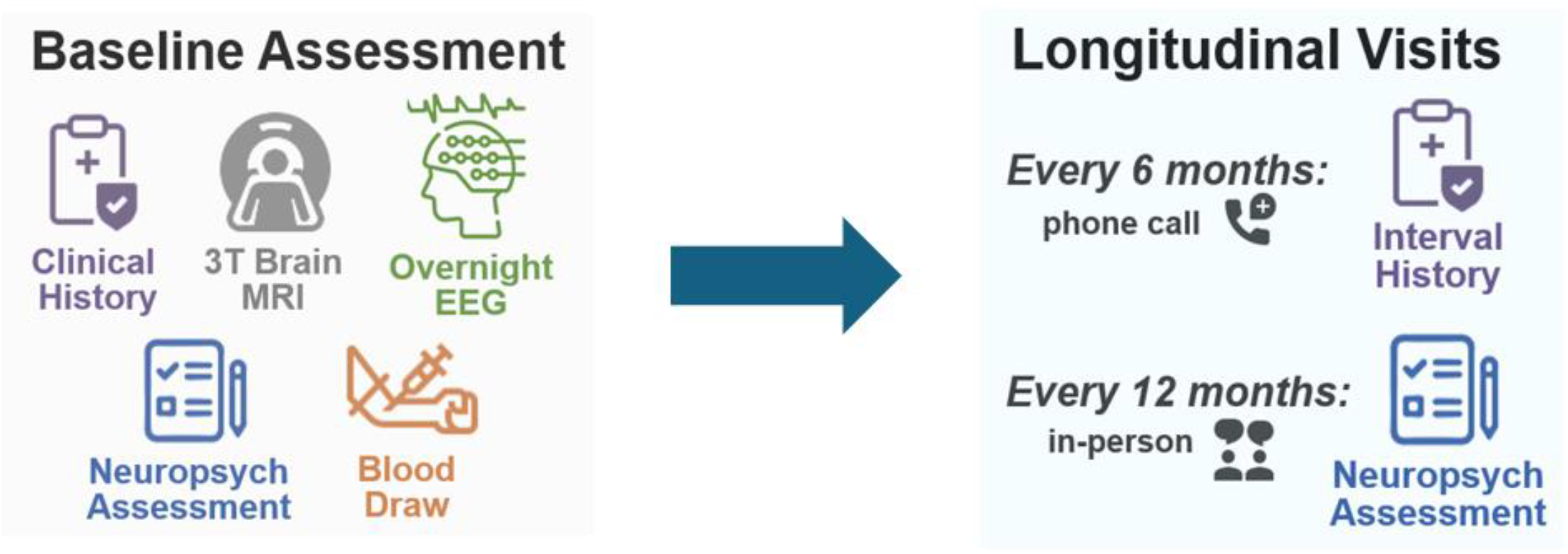
Schematic of ELUCID Study Protocol.

#### Baseline Assessment

We will obtain a comprehensive clinical history focused on seizure history; medical comorbidities; medications; risk factors for dementia, cardiovascular disease, and autoimmune disease; family history; and social history. Participants will undergo a 3T Brain MRI, at-home overnight scalp EEG, blood draw, and baseline neuropsychological assessment

The Clinical Intake Questionnaire includes questions on:

- Demographics
- Epilepsy history and risk factors, including family history
- Dementia risk factors, including family history
- Auto-immune disease history and risk factors, including family history
- Medical history
- STOP-BANG questionnaire for obstructive sleep apnea
- FRAIL scale questionnaire for frailty
- Lifestyle factors, including education, occupation, diet, physical activity, and substance use
- Social determinants of health

The Interval History Questionnaire assesses interval changes over the prior 6 months, for the following:

- Seizure frequency and severity
- Changes to anti-seizure medications and other prescription medications
- Subjective changes in cognition / memory
- Changes in medical history
- Changes in living situation
- Development of primary outcomes of interest: mild cognitive impairment, dementia
- Development of secondary outcomes of interest: ischemic or hemorrhagic stroke, transient ischemic attack, myocardial infarction, serious cardiac arrhythmia, and mortality

Annual neuropsychological assessments include the following tests and questionnaires:

- Mini-Mental State Exam
- Digit symbol substitution test
- Trail making test
- Category fluency (animals, vegetables, fruits)
- Phonemic fluency (F, A, S words)
- Logical Memory delayed story recall
- Free and Cued Selective Reminding Test
- Boston Remote Assessment for Neuro-cognitive Health^41^
- Geriatric Depression Scale
- Quality of Life in Epilepsy Scale - 31 item questionnaire
- Quick Dementia Rating Scale (QDRS, completed by both participant and study partner)
- Cognitive Function Instrument (CFI, completed by both participant and study partner)
- PACC-5 composite (derived from MMSE, DSST, Logical memory, FCSRT, category fluency)

Physical exam: Includes measurements of height, weight, neck circumference, waist circumference, blood pressure, and heart rate.

3T Brain MRI: Research MRIs are acquired on a Siemens Prisma 3T MRI scanner. The ELUCID MRI protocol is closely aligned with the Alzheimer’s Disease Neuroimaging Initiative (ADNI-3 Basic) MRI protocol, to facilitate comparison to ADNI-3 data and use of ADNI-3-derived, age- and sex-matched non-epilepsy control groups. Core MRI sequences that will be acquired include: Sagittal 3D Accelerated MPRAGE; Sagittal 3D FLAIR; Accelerated High Resolution Hippocampus Scan; and Axial T2 Star. When possible, the following sequences will also be acquired: Axial DTI; Axial 3D pASL; and Axial resting state MRI (10 minutes, eyes open).

Overnight scalp EEG: Registered EEG technologists will set up scalp EEGs in participants’ homes, using the International 10-20 system with additional T1/T2 electrodes and a single-lead EKG. EEGs will be recorded on a 24-channel Lifelines device, sampled at 200 Hz.

Fasting blood draw: To ensure uniformity across sites, materials for blood collection and processing at each site are coordinated through BioSEND, an NIH-sponsored biorepository service. Approximately 30cc blood will be collected by venipuncture into 2 × 10mL lavender top EDTA tubes, 1 × 3mL lavender top EDTA tube, and 1 × 4.5mL green top LiHep plasma separator tube. *For plasma and buffy coat*: within 30 minutes of collection, samples from the 2 × 10mL EDTA tubes are centrifuged at 1500xg at 4°C for 15 minutes. Plasma and buffy coat layers are drawn off, aliquoted into pre-chilled labeled cryotubes, and frozen at -80°C within 2 hours of collection. *For serum*: within 90 minutes of collection, samples from the 1 × 4.5mL LiHep tube are centrifuged at room temperature at 1500xg for 15 minutes. Serum is drawn off, aliquotted into labeled criovials and frozen at -80°C within 2 hours of collection. *For whole blood*: samples from the 1 × 3mL EDTA tube are frozen at - 80°C within 2 hours of collection. Processed samples from all participants are shipped on dry ice to the BioSEND Repository, for long-term storage and management.

#### Phone Follow-Up Visits

Phone Visits include the Interval History Questionnaire, as described above in the

*Baseline Assessment*.

#### In-Person Follow-Up Visits

In-person Visits include the Interval History Questionnaire, Physical Exam, and Neuropsychological Assessment, as described above in the *Baseline Assessment*.

### Data Collection and Management

Data from questionnaires, physical examination, and neuropsychological assessments are stored in a central Research Electronic Data Capture (REDCap) database. The data and neuropsychological test scores are input into REDCap by research coordinators and are reviewed and validated by 2 independent reviewers for accuracy and completeness.

MRI data are stored centrally and reviewed regularly for quality control purposes. Cortical reconstruction and subcortical segmentation are performed using FreeSurfer. Cortical thickness and subcortical volumes are extracted from MP-RAGE images. Automated WMH segmentation is performed on T2 FLAIR images, and

WMH volumes are calculated, normalized to intracranial volume.

EEG data are stored centrally and reviewed regularly for quality control purposes. Board-certified epileptologists / clinical neurophysiologists will visually review the EEGs to determine the location and severity of EEG slowing and epileptiform abnormalities. Computational algorithms previously developed by our team will be used to extract quantitative features from the EEGs, including automated spike detection^42,43^ and extraction of sleep macro-architectural and micro-architectural features^44,45^.

Blood samples are shipped to BioSEND for long-term storage and management. All samples undergo quality control checks at BioSEND to assess for hemolysis, inadequate volume, and other nonconformities. DNA is extracted from the buffy coat sample, and a genetic fingerprint is obtained, yielding APOE genotype and LRRK2 G2019S mutation status. The following blood biomarkers will also be obtained:

- General health and vascular risk: creatinine, fasting lipid panel, and hemoglobin A1C.
- Neuropathology: AB42, AB40, p-tau217, NfL, GFAP
- Inflammatory markers: IFN-g, IL-1b, IL-2, IL-4, IL-6, IL-10, IL-12p70, IL-17A, and TNF-a
- Vascular health markers: bFGF, Flt1, PlGF, VEGF, VEGF-C, VEGF-D

### Outcome measures

Our primary outcomes of interest are development of MCI and dementia. As indicated above, individuals with MCI are eligible for enrollment in the study, and in these participants, we will assess the primary outcome of dementia. We will assess development of MCI and dementia at each study visit using NIA-AA criteria^46,47^, implemented as shown in **Table 3**. Participants who meet criteria for MCI or dementia will be discussed at regularly scheduled consensus meetings with the ELUCID study behavioral neurologist and neuropsychologist to adjudicate their diagnoses. If the consensus is that a participant meets criteria for MCI or dementia, the Site PI will inform the participant and their epileptologist of the diagnosis and recommend that the participant undergo further clinical evaluation for this.

**Table 3.** Implementation of NIA-AA Criteria for Diagnosis of MCI and Dementia.

| **MCI** | **Dementia** |
| --- | --- |
| • Subjective concern for cognitive decline, as reported by participant [or partner], based on: "Compared to 1 year ago, do you feel that you have [the participant has] had a significant decline in memory?"<br>• MMSE score 24-30, inclusive<br>• Objective impairment in $\geq 1$ cognitive domain (score on any test $\geq 1.5$ SD below age- and education-matched norm)<br>• Preserved global functioning (QDRS 2-5, inclusive) | • MMSE score $\leq 26$<br>• Objective impairment in $\geq 2$ cognitive or behavioral domains (2+ test scores $\geq 1.5$ SD below age- and education-matched norms, across different domains)<br>• Impaired global functioning (QDRS $\geq 6$) |

Our secondary outcomes of interest are development of incident stroke (ischemic or hemorrhagic), transient ischemic attack, myocardial infarction, serious cardiac arrhythmia, and death.

## DATA ANALYSIS PLAN & STATISTICAL CONSIDERATIONS

### Aim 1: Establish an unbiased framework for identifying mechanisms of dementia in LoUE

We will use unsupervised machine learning approaches to organize LoUE into disease subtypes based on pathology, clinical phenotype, and cognitive trajectory.

(a) Subtypes based on pathology. We will identify individuals with AD pathology, vascular risk and pathology, inflammatory profiles, and mixed pathologies, based on blood and imaging biomarkers and medical comorbidities. We will estimate the prevalence of each pathology and the fraction of LoUE without these pathologies (“Undifferentiated” subtype).

(b) Subtypes based on clinical phenotype. We will use a data-driven, unsupervised machine learning (UML) approach to phenotype discovery. Phenotypes will be derived from combinations of clinical and diagnostic features in **Tables 4 and 5**. We will develop 2 kinds of phenotypes: 1) <u>Basic</u>: uses only features readily available to clinicians (**Table 4**), allowing ease of interpretation and translation to clinical use; and 2) <u>Advanced</u>: uses all basic features in **Table 4**, combined with advanced features in **Table 5**, allowing for more comprehensive and nuanced phenotypes.

**Table 4.** Basic clinical features for analysis.

| Demographics | Medical Co-morbidities | Lifestyle | Family History | Epilepsy features |
| --- | --- | --- | --- | --- |
| - Age at seizure onset<br>- Sex<br>- Race<br>- Ethnicity<br>- Education level<br>- Zip code | - Hypertension<br>- Hyperlipidemia<br>- Diabetes<br>- Coronary artery disease<br>- Peripheral vascular disease<br>- Congestive heart failure<br>- Obstructive sleep apnea<br>- Cancer<br>- Chronic lung disease<br>- Renal or hepatic disease<br>- Autoimmune disease<br>- Psychiatric disease | - Tobacco history<br>- Alcohol history<br>- Recreational drug use<br>- Physical activity level<br>- Dietary habits | - Epilepsy<br>- Dementia<br>- Vascular disease<br>- Autoimmune disease<br>- Cancer | - Seizure semiology<br>- Convulsive seizures<br>- Nocturnal seizures<br>- Seizure duration<br>- Seizure frequency<br>- Epilepsy risk factors<br>- Seizure medications<br>- Medication response<br>- QOLIE-31 score |

**Table 5.**
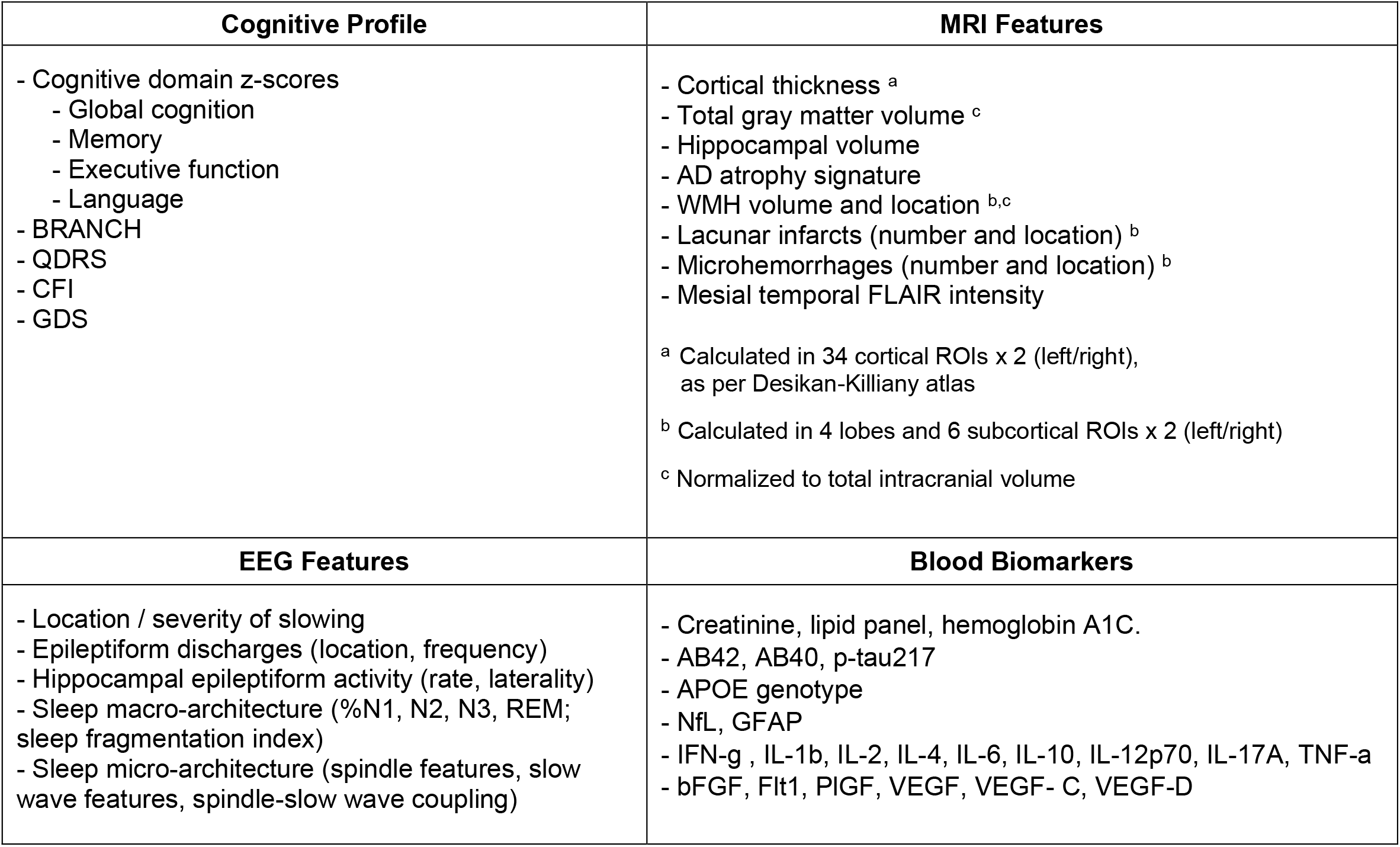
Advanced features for analysis.

| Cognitive Profile | MRI Features |
| --- | --- |
| - Cognitive domain z-scores<br>- Global cognition<br>- Memory<br>- Executive function<br>- Language<br>- BRANCH<br>- QDRS<br>- CFI<br>- GDS | - Cortical thickness <sup>a</sup><br>- Total gray matter volume <sup>c</sup><br>- Hippocampal volume<br>- AD atrophy signature<br>- WMH volume and location <sup>b,c</sup><br>- Lacunar infarcts (number and location) <sup>b</sup><br>- Microhemorrhages (number and location) <sup>b</sup><br>- Mesial temporal FLAIR intensity<br><sup>a</sup> Calculated in 34 cortical ROIs x 2 (left/right), as per Desikan-Killiany atlas<br><sup>b</sup> Calculated in 4 lobes and 6 subcortical ROIs x 2 (left/right)<br><sup>c</sup> Normalized to total intracranial volume |
| EEG Features | Blood Biomarkers |
| - Location / severity of slowing<br>- Epileptiform discharges (location, frequency)<br>- Hippocampal epileptiform activity (rate, laterality)<br>- Sleep macro-architecture (%N1, N2, N3, REM; sleep fragmentation index)<br>- Sleep micro-architecture (spindle features, slow wave features, spindle-slow wave coupling) | - Creatinine, lipid panel, hemoglobin A1C.<br>- AB42, AB40, p-tau217<br>- APOE genotype<br>- NfL, GFAP<br>- IFN-g , IL-1b, IL-2, IL-4, IL-6, IL-10, IL-12p70, IL-17A, TNF-a<br>- bFGF, Flt1, PIGF, VEGF, VEGF- C, VEGF-D |

We will use 2 UML methods: agglomerative hierarchical clustering (AHC) and non-negative tensor factorization (NNTF) to identify clinical phenotypes. We will evaluate each approach and determine the best stopping criterion based on metrics of intrinsic clusterability, cluster distinctness, and cluster reproducibility. The biological relevance of each identified phenotype in terms of its relevance for predicting cognitive outcomes, will be assessed in Aims 2 and 3.

(c) Subtypes based on cognitive trajectory. We will identify distinct cognitive trajectories in LoUE, using baseline cognitive performance and longitudinal change in performance across multiple cognitive domains. Initially, for ease of interpretation, we will stratify participants into the cognitive trajectory subtypes using measures of global cognition (PACC-5) and z-score domain composites for memory, language, and executive function. z-score composites will be calculated using age- and sex-based norms for each test and averaging an individual’s z-scores across tests in each domain, and at each time point. Impairment will be defined as a z-score composite ≤ -1.5 in any domain. Cognitive decline will be defined as development of impairment (z-score composite ≤ -1.5) in a previously cognitively normal participant, or a decrease from z-score composite baseline of ≥ 1 in a previously impaired participant. We will determine the prevalence of each trajectory and the affected domains. *Data-driven approach:* We will also model cognitive trajectories using multivariable group-based trajectory modeling (MGBTM), an UML method for discovering clusters within longitudinal data. MGBTM is a finite mixture model that assumes a population is composed of subgroups that follow distinct trajectories of one or more repeated measures over time, e.g., cognitive test scores. MGBTM uses a data-driven approach to probabilistically cluster individuals who follow similar trajectories^48–50^. MGBTM models will include baseline and longitudinal data from all neuropsychological tests. To select the optimal number of trajectories, we will use Bayesian Information Criteria to balance model fit and parsimony.

### Aim 2. Identify clinical features and biomarkers associated with development of cognitive decline and dementia in LoUE

Most individuals with LoUE do not develop dementia within 3-4 years of onset. We will examine a wide array of features, described below, and identify those associated with cognitive decline and dementia in LoUE, by comparing differences between the following LoUE subgroups: 1) LoUE with normal vs. impaired cognition at baseline; 2) LoUE with stable vs. declining cognition at 3-year follow up; 3) LoUE with normal cognition vs. dementia at 3-year follow up; and 4) Cognitive trajectory sub-types of LoUE, as identified in Aim 1.

1. Epilepsy and treatment-related features: Epilepsy severity will be assessed by (a) 1-year terminal seizure remission (seizure-freedom for ≥ 1 year prior to last follow-up); and (b) duration of active epilepsy (time between first seizure and achieving 1-year terminal remission, or until last follow-up if remission is not achieved). Epilepsy location will be categorized as dominant vs non-dominant hemisphere, and temporal vs extratemporal location. Epilepsy medications will also be assessed, as well as use of anti-hypertensive or anti-platelet medications.
2. Blood biomarkers: We will examine the blood-based biomarkers in **Table 5**, which span AD, vascular, and inflammatory contributions to cognitive decline.
3. Neurophysiologic features: We will examine scalp EEG features in **Table 5**, which span epileptiform activity and sleep physiology
4. Structural features: We will assess region-of-interest (ROI)-based MRI features (**Table 5**). In addition to identifying MRI features associated with cognitive decline in LoUE, we will also identify MRI features associated with LoUE, by comparing our LoUE cohort to a control group without epilepsy, derived from the ADNI-3 cohort^51,52^. The control group will consist of 200 ADNI-3 participants, matched to our LoUE cohort by age, sex, race, and baseline cognitive status. ADNI-3 participants were between ages 55-90 at enrollment, and seizures were an exclusion criterion. MRIs acquired in ADNI-3 used standard acquisition protocols, which the ELUCID MRI protocol closely matches. ADNI-3 MRIs will be processed using the same pipelines used for ELUCID MRIs. Identifying MRI features associated with cognitive decline in LoUE that are distinct from features associated with LoUE alone would provide evidence that mechanisms underlying seizures and cognitive decline are separable.

Analysis: We will first assess group differences using 2-sided t-tests (continuous features) or Chi-square tests (dichotomous features). As a conservative estimate, with 600 LoUE and 200 ADNI-3 controls, and a Bonferroni-corrected alpha of 0.001 or 0.0001 (50 to 500 comparisons), we will have 90% power to reject the null hypothesis of zero effect size when the population effect size (Cohen’s d) is 0.37 or 0.42, respectively (low to medium effect sizes). For features with significant group differences at the univariate level, we will perform adjusted analyses using multinomial logistic regression, where LoUE subgroup is the dependent variable, and each feature is the primary predictor, with co-variates of age, sex, race, education level, and APOE ε4.

### Aim 3. Develop practical clinical tools to forecast an individual’s risk of developing mild cognitive impairment and/or dementia after presentation with LoUE

1. Prognostication based on LoUE subtypes: We will evaluate whether LoUE subtypes (Aim 1), can be used to predict cognitive outcomes. To test whether a LoUE subtype predicts cognitive decline, we will use linear mixed effects models with random intercept and slope, where the dependent variable is the PACC-5, QDRS, or domain z-score composite. The primary predictor will be LoUE subtype (fixed effect). We will adjust for time in study (as a stand-alone variable, and as an interaction between etiology and time; fixed effects). We will control for covariates (age, sex, race, education level, epilepsy duration, epilepsy severity, number and type of seizure medications, use of anti-hypertensive and anti-platelet medications, and APOEε4 allele number (fixed effects)); and for random effects (intercepts for individual participants and time-by-participant random effects). To test whether a LoUE subtype predicts outcomes of MCI or dementia, we will use Cox proportional hazards (PH) models adjusted for death as a competing risk^53,54^, with date of seizure onset as the origin, date of MCI or dementia onset as the event time, and LoUE subtype as the independent variable. Censoring will occur at last follow-up. Covariates will include age, sex, race, education, seizure frequency, number of seizure medications, use of anti-hypertensive and anti-platelet medications, and APOE ε4 allele number. Some participants may develop incident stroke, which can affect cognition. If needed, we will adjust for stroke as an additional competing risk. To test whether a LoUE subtype predicts cognitive trajectory, we will use multivariate multinomial logistic regression, with LoUE subtype as independent variable, covariates as above, and cognitive trajectory subtypes (Aim 1) as target outcomes.
2. Developing tools to forecast risk of dementia: We will use multivariable Cox regression to develop a prognostic model for risk of MCI or dementia within 3 years following a diagnosis of LoUE. As in Aim 1, we will develop a basic model (using only features available to clinicians (**Table 4**), medications, and basic LoUE phenotypes (Aim 1)) and an advanced model (using features from **Tables 4 and 5**, medications, and advanced LoUE phenotypes (Aim 1)). Lasso regularization will be used to select features and prevent overfitting. Performance will be assessed using 5-fold cross-validation. The Cox model will estimate how well baseline clinical features of LoUE can predict future MCI or dementia, but will be too complex for clinical use. To reduce this to a simple, interpretable, point-based tool to predict a LoUE patient’s risk of dementia or MCI within 3 years, we will use Risk-calibrated Supersparse Linear Integer Model (RiskSLIM)^55^. Unlike black-box machine learning models, RiskSLIM produces simple yet accurate models with “points” that are added to give a final risk score. The forecasting models will be evaluated using Harrel’s c-statistics calculated via 10-fold cross validation^56^.

### Statistical considerations

Our primary outcomes are development of incident MCI and dementia. Total follow-up time, from first seizure to end of study, will range from 2 years (enrolled in Year 3, first seizure that year) to 8 years (enrolled in Year 1, with first seizure 3 years prior). Prior small studies of LoUE have shown that 50-60% of individuals with LoUE have MCI at the time of their first seizure^57,58^. We conservatively estimate that 40-50% of our study population will have MCI on enrollment in the study. We estimate that 25% of participants will develop incident dementia during the study^30,31,33,59^. A two-sided logrank test with 600 LoUE participants and 25% in a subtype of interest achieves 80% power at a 0.0125 significance level to detect a hazard ratio of 1.84 if the control group (all other participants) hazard rate is 0.055 or lower. Even if only 10% of participants develop dementia, we will be powered to detect a hazard ratio of 2.62. Placed in context, a small prospective study found that 37.5% of LoUE had AD pathology (pathologic CSF Aβ1-42), with a hazard ratio of 3.4 for progression to AD dementia^59^. These considerations suggest that our sample provides sufficient power to characterize the risks of MCI and dementia associated with LoUE subtypes.

## DISCUSSION

Here we present the rationale and design for the ELUCID study, whose over-arching goal is to advance understanding of the mechanisms underlying MCI and dementia in LoUE, and to develop tools to predict which individuals with LoUE are at greatest risk for developing MCI and dementia. ELUCID will generate a large, clinically annotated, multi-modal biorepository for LoUE research that will be made publicly available for further advancement of LoUE research. Moreover, we will use data-driven methods to identify clinically relevant LoUE subtypes that can serve as a starting point for developing a precision medicine approach to preventing dementia and other adverse outcomes in LoUE.

While individuals with LoUE comprise a large and growing population at high risk for AD/ADRD, a history of epilepsy has been an exclusion criterion in almost every AD clinical trial to date. As new therapies for AD/ADRD are approved, individuals with epilepsy will be largely excluded even if they have underlying AD^60^ or other targeted pathology. ELUCID will lay the groundwork and develop tools needed to carry out informative and efficient clinical trials in LoUE. In the future, our hope is to expand AD/ADRD therapeutic development to the LoUE population and explore unique and/or combined treatment approaches (e.g., anti-amyloid therapies with network modulation) to reduce dementia risk in LoUE.

## Data Availability

This is a protocol paper for a prospective study. There are no data produced in the present work.

## AUTHOR CONTRIBUTIONS

Protocol design: ADL, MBW, ELJ, RAS, NJ, LBC, DNG, REA, GAM Drafting of manuscript: ADL, MBW Revising of manuscript: ADL, MBW, REA, LJB, LBC, TG, NJ, DNG, ELJ, GAM, RAS, MMS, RZ

